# Real-world usage diminishes validity of Artificial Intelligence tools

**DOI:** 10.1101/2022.11.17.22282440

**Authors:** Akhil Vaid, Ashwin Sawant, Mayte Suarez-Farinas, Juhee Lee, Sanjeev Kaul, Patricia Kovatch, Robert Freeman, Joy Jiang, Pushkala Jayaraman, Zahi Fayad, Edgar Argulian, Stamatios Lerakis, Alexander Charney, Fei Wang, Matthew Levin, Benjamin Glicksberg, Jagat Narula, Ira Hofer, Karandeep Singh, Girish N Nadkarni

## Abstract

**Background:** Substantial effort has been directed towards demonstrating use cases of Artificial Intelligence in healthcare, yet limited evidence exists about the long-term viability and consequences of machine learning model deployment.

**Methods:** We use data from 130,000 patients spread across two large hospital systems to create a simulation framework for emulating real-world deployment of machine learning models. We consider interactions resulting from models being re-trained to improve performance or correct degradation, model deployment with respect to future model development, and simultaneous deployment of multiple models. We simulate possible combinations of deployment conditions, degree of physician adherence to model predictions, and the effectiveness of these predictions.

**Results:** Model performance shows a severe decline following re-training even when overall model use and effectiveness is relatively low. Further, the deployment of any model erodes the validity of labels for outcomes linked on a pathophysiological basis, thereby resulting in loss of performance for future models. In either case, mitigations applied to offset loss of performance are not fully corrective. Finally, the randomness inherent to a system with multiple deployed models increases exponentially with adherence to model predictions.

**Conclusions:** Our results indicate that model use precipitates interactions that damage the validity of deployed models, and of models developed in the future. Without mechanisms which track the implementation of model predictions, the true effect of model deployment on clinical care may be unmeasurable, and lead to patient data tainted by model use being permanently archived within the Electronic Healthcare Record.

## Introduction

Artificial Intelligence (AI) promises to bring transformative change to the practice of medicine^1^. Underpinning this promise are machine learning (ML) models – mathematical constructs capable of deriving patterns within data. The creation of performant, generalizable models is heavily dependent on access to copious, high-quality data^2^. In healthcare settings, the Electronic Healthcare Record (EHR) is the primary mechanism for storage, retention, and retrieval of patient data^3^; making it the principal source of truth both for model inputs, and *labels* indicative of the occurrence and timing of clinical outcomes. As such, ML models are inexorably linked to the EHR during the entirety of their lifecycle i.e., during development, and post-deployment.

Substantial effort directed towards the early, developmental part of this process has translated into a large body of work demonstrating use cases of ML in healthcare^4-7^, and commentary about best practices regarding model development^8-10^. Conversely, relatively little attention has been paid to the later, post-deployment part of the process^11^. This is concerning since post-deployment monitoring has revealed that performance in laboratory settings does not guarantee success in real-world settings^12^. Real-world usability of ML models is entirely contingent upon on the maintenance on the data and label relationship^13^, and model failure manifesting as unacceptably high error rates may occur when data encountered during deployment is substantially different from data available during development. For example, the COVID-19 pandemic resulted in a *dataset shift* leading to the decommissioning of a sepsis detection model^14^. The recommended solution for this problem is *re-training* and re-calibrating the model against a larger corpus of patient data representative of all patient groups^14^. However, since this solution does not consider the relationship between models and the EHR, it may exacerbate an insidious phenomenon. Paradoxically, this is the effect of a model working as intended.

ML in healthcare is tied to one central thesis – a deployed model will help improve patient outcomes^15^. This can only occur if model predictions modify both provider behavior, and the natural course of disease beyond what is seen in usual medical practice. However, since such modifications are recorded within the EHR, deployed models have the ability to interact with the apparatus of their creation. Mechanistically, a model recognizing a pattern associated with a deleterious clinical outcome (e.g., death) will signal the need for more intensive monitoring or alternative care pathways. Ideally, implementation of these measures will prevent the outcome, essentially *inverting* the label that the model predicted. Simultaneously, both the pattern at presentation, and lack of an outcome will come to be associated with that patient’s records in perpetuity. Extending this to the entire EHR, while most hazardous patterns will continue to correspond to the outcome, an increasing number of these patterns will start to correspond to the outcome not occurring. These mixed associations now form part of any new data sourced from the EHR, potentially confounding any attempts at re-training the model.

Further, the sequalae and underlying pathology of a deleterious clinical event can extend to involve other organ systems within the body, and precipitate linked clinical events. For example, either of acute kidney injury (AKI) or sepsis can be a precursor to mortality^16,17^. Within such *clinical cascades*, preventing or delaying the incidence of an upstream event can offset the incidence of the downstream outcome^18^. Therefore, a model able to prevent the initial event will culminate in the EHR recording a hazardous pattern at presentation, and non-occurrence of both the initial and linked outcome. As before, these mixed associations may confound the development of any future model for the linked outcome.

Finally, models are trained on and make predictions on the *biological state* of a patient as described by their labs, vitals, and imaging. More predictions are available for each patient with multiple model deployment. Upon implementation, any actions taken due to a prediction may influence biological state sufficiently to render an existing prediction from another model invalid or inapplicable.

In each case, model implementation precipitates *interactions* wherein usage of one model influences the function of another. Such interactions are unintended, and their effects largely unaccounted for given the relative novelty of real-world machine learning model deployments in healthcare settings. To better understand these effects, we create a framework that simulates model deployment in addition subsequent changes in clinical outcomes and model performance using real-world data from a diverse cohort of New York City patients, and a publicly available dataset of ICU admissions^19^. We discuss the implications of these interactions on the future of machine learning in healthcare and recommend measures to mitigate their effects.

## Methods

### Data sources and outcomes

We utilize data from two sources – the MIMIC IV database^19^ containing records for inpatient ICU admissions to the Beth Israel Deaconess medical center from 2008 – 2019; and pooled data from five facilities within the Mount Sinai Health System (MSHS) – containing inpatient ICU admission records from Jan 2010 – Jan 2020. For both sources, we extract demographics, vitals, and lab investigations restricted to a period of 12 hours prior to an “anchor” time related to the outcome of interest. We model common clinical outcomes, i.e. AKI^20^ in critically ill patients, critical care mortality, and post cardiac catheterization AKI **(Supplementary Table 1)**. Salient characteristics of patient populations for each outcome of interest are shown in **Supplementary Tables 2 – 4**.

The study was approved by the Institutional Review Board at the Icahn School of Medicine at Mount Sinai. The study was exempt from the requirement of informed consent owing to its retrospective design.

### Model development and evaluation

We train and evaluate both complex non-linear models (XGBoost^21^), and simpler, explainable linear models (LASSO) for outcomes of interest.

Model performance^22^ is measured using threshold independent metrics i.e., Area Under the Receiver Operating Characteristic Curve (AUROC), and Area Under the Precision Recall Curve (AUPRC), in addition to threshold dependent metrics i.e., sensitivity, specificity, and accuracy. **(Supplementary Methods)**

### Label inversion

We define a *label inversion* as a change in recorded outcome due to the influence of a deployed model. **(Figure 1A.)**

**Figure 1:**
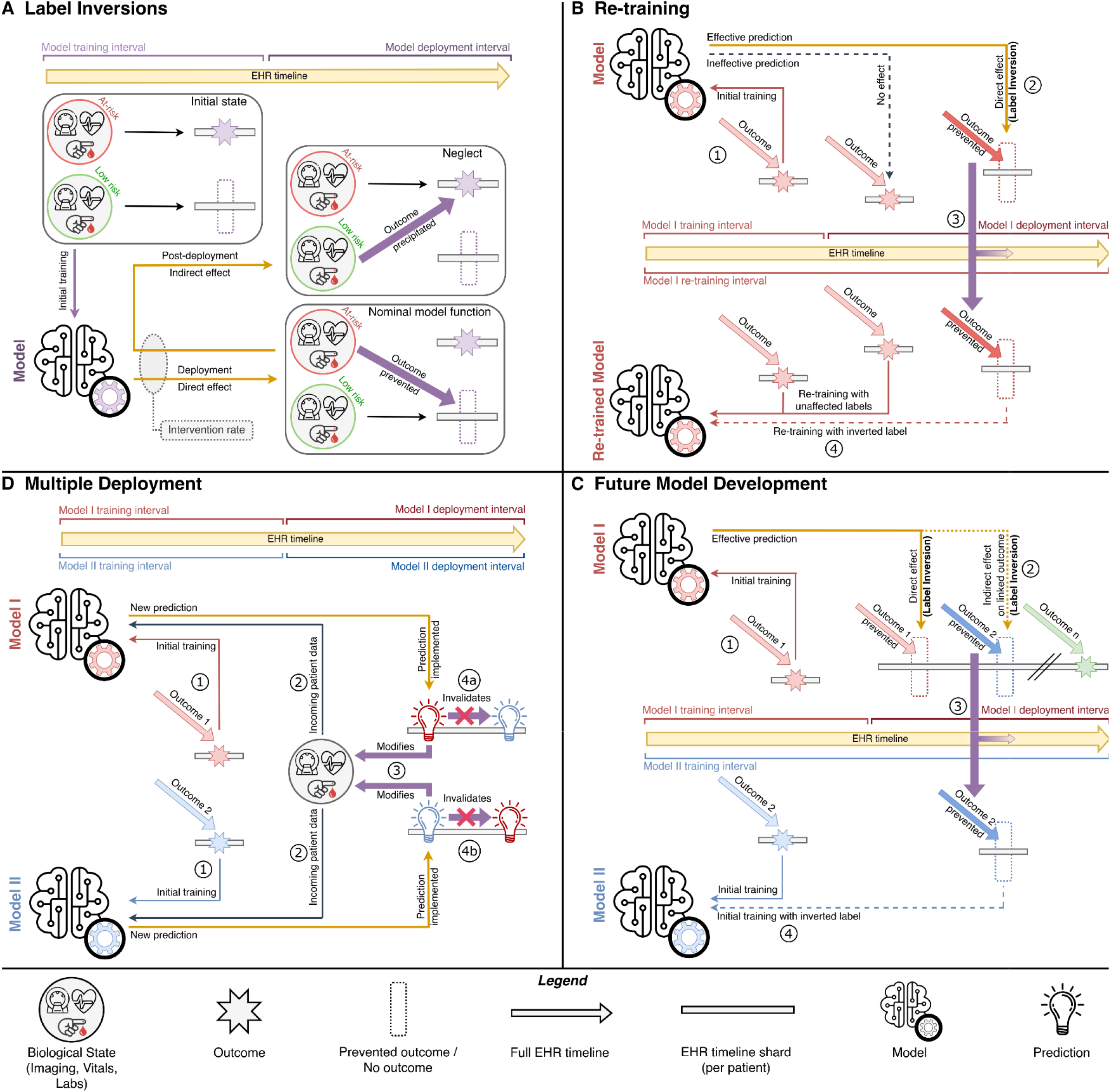
Overview of label inversions and simulation scenarios. Panel A shows a schematic of how label inversions occur, both in the case of nominal model function, and neglect caused by over-reliance on a model, or redistribution of clinical resources due to a model. Panel B shows an overview of Scenario I. A model is trained as usual (1), and its use leads to prevention of the deleterious outcome it was trained to detect (2) – leading to a label inversion. The inverted label is inserted into the EHR, as well as the training dataset at the time of re-training (3), followed by the actual re-training of the model with the inverted label in place (4). Panel C shows an overview of Scenario II. A model designed to prevent a deleterious clinical outcome (1) prevents both that outcome, as well as a downstream linked outcome – leading to a label inversion (2). The inverted label is inserted into the EHR, as well as the training dataset for a future model developed for the linked outcome (3), followed by the development of a model for the linked outcome (4). Panel D shows an overview of Scenario III. Models for two separate outcomes are trained independently (1) and deployed simultaneously. With incoming data on the biological state of new patients (2), predictions are issued together. The first implemented prediction modifies this biological state (3), resulting in other predictions being rendered invalid (4a, 4b).

### Quantification of model usage and effectiveness

We define *intervention rate* (IR) as the proportion of true positive model predictions adhered to leading to changes in management – independent of effect on outcome. We also define a separate *effective intervention rate (EIR)* as the proportion of true positive model predictions that are both adhered to by healthcare workers and induce a label inversion.

Either rate is given as a number between 0 – 1, and simulated as discrete values of 0, 0.05, 0.1, 0.2, 0.5, and 0.75. A value of 0 is equivalent to model development in laboratory conditions and implies the model had no real-world impact. Higher values imply greater application and/or effectiveness of predictions. For example, an intervention rate of 0.1 implies 10% of true positive predictions led to a change in care, whereas an effective intervention rate of 0.1 implies 10% of true positive predictions prevented a deleterious outcome.

### Simulation scenarios

We consider three scenarios of model deployment detailed algorithmically in **Supplementary Methods**. Scenarios I and II consist of simulations examining re-training and future model development respectively at different EIR’s. Scenario III examines multiple, simultaneous model deployments at different IR’s.

For Scenario I (*Re-training*), each problem-specific dataset is partitioned into ten intervals of equal duration. We train a model given data for an interval and simulate its deployment during the next interval by inducing label inversions according to EIR. The model is re-trained, recalibrated, and its performance evaluated at the end of each such interval keeping prior label inversions in place. **(Figure 1B.)**

For Scenario II (*Future model development*), each dataset is partitioned into a development and deployment interval. Using data from the development interval, we train a model for an outcome from earlier in a clinical cascade. Label inversions according to EIR are induced for a linked outcome within data from the deployment interval. Following this, we develop a model for the linked outcome keeping label inversions in place. **(Figure 1C.)**

For Scenario III (*Multiple deployment*), we simulate concurrent model deployment. Absent protocol or centralized documentation about other models in play, model predictions are implemented in random order as influenced by IR. Implemented predictions modify baseline biological state on which predictions were made and render other/prior predictions invalid for use. **(Figure 1D.)** Effective accuracy is calculated as the ratio of the number of valid predictions matching the target outcome vs the total number of predictions.

### Mitigations

We consider two dataset modification methods to mitigate the effect of label inversions for scenarios I and II. One, the addition of a feature that keeps track of whether a patient has had a prediction implemented. Two, dropping those patients from the dataset for whom predictions have been implemented. **(Table 1)**

**Table 1.** Mitigation measures for each deployment scenario

|  | Mitigation | Description | Caveats |
| --- | --- | --- | --- |
| Scenarios I and II | Keep track of all implemented predictions | Mechanisms which deliver model predictions to clinicians also keep track of whether a physician decides to implement these predictions. | Will require immediate cessation of model use until better prediction delivery mechanisms can be implemented. |
|  | Additional feature | Utilize an additional feature that makes a model aware if a patient has had a prediction implemented before. | Simulations show models tend to overfit to the feature and prospective testing performance falls for re-training.<br>Image/text-based models will need to be restructured to account for an additional tabular feature. |
|  | Remove patients with implemented predictions from dataset | Drop all data points with an implemented positive prediction for re-training the same model, or for development of any new model. | Exacerbation of class imbalance in clinical datasets.<sup>36</sup><br>Experiments have been performed with removals in proportion to intervention rate. Unless accurate record keeping is performed, all patients with a positive prediction may be rendered unsuitable for future inclusion in training data. |
|  | Model selection | Optimized XGBoost models outperform optimized LASSO models for the same datasets, subject to the same constraints. | Less transparent/Black-box models may reduce clinician acceptance.<sup>37</sup> |
| Scenario III | Restrict number of models applied to one patient | Selective use of models. | Reduces usability of ML models. |
|  | Time between prediction and implementation | Reduce the time a prediction is “valid” for. E.g., 12-hour mortality risk instead of 3-day mortality risk. Reduces chances of interference from another model. | Datasets may become extremely imbalanced<sup>36</sup> since not all outcomes are proximal to the time of data collection. |
|  | Reissue predictions | Predictions from *all* models relevant to a patient must be reissued after a change in management. Predictions from models which influence outpatient care must be refreshed regularly especially if they influence long term therapy. | Expensive to continuously collect new model specific data for prediction pipelines and deliver resulting predictions. |

### Neglect

ML models may precipitate patient neglect due to provider adherence to false negative model predictions resulting in incorrect patient triage^11^ **(Figure 1A, Supplementary Table 5)**. We quantify this effect by inducing label inversions for such predictions. For Scenario I, we induce inversions for true negatives, and for Scenario II, we induce inversions in the linked outcome following a false negative prediction for the initial outcome. In either case, we limit the degree of neglect to 1% of such predictions independently of the EIR. Neglect is not factored in for Scenario III since unimplemented negative predictions cannot influence patient state.

## Results

We perform multiple in-silico experiments within each scenario accounting for variations in deployment conditions **(Supplementary Table 5)**. Changes in model performance are measured at different intervention rates and compared to baseline performance with an intervention rate of 0.

The following results describe simulation experiments reflective of real-world conditions – classification thresholds are set for a sensitivity of 90%, population class distribution is maintained, hyperparameter optimization is performed, neglect is factored in for scenarios I and II, and no mitigations are in place.

### Scenario I – Re-training

We simulate the deployment of a model for prediction of post-ICU admission mortality in a large, tertiary care hospital center using data from the MIMIC IV database.

We observe a loss of performance across all metrics after one or two iterations of re-training followed by a relative stabilization. Compared to baseline, AUROC drops 9.5% (0.82 to 0.74), AUPRC drops 15.8% (0.38 to 0.32), and specificity drops 38.6% (0.44 to 0.27) at the lowest simulated EIR of 0.05 after one iteration of re-training. **(Figure 2, Supplementary Table 6)** Cumulative label inversions and loss in performance increase with increasing EIR. Average specificity across all iterations of re-training drops 20% (0.60 to 0.48) over baseline at an EIR of 0.05, and 31.1% at an EIR of 0.5 (0.60 to 0.41). Applicable mitigations mostly correct AUPRC, but not loss in specificity. **(Supplementary Figure 1)**

**Figure 2.**
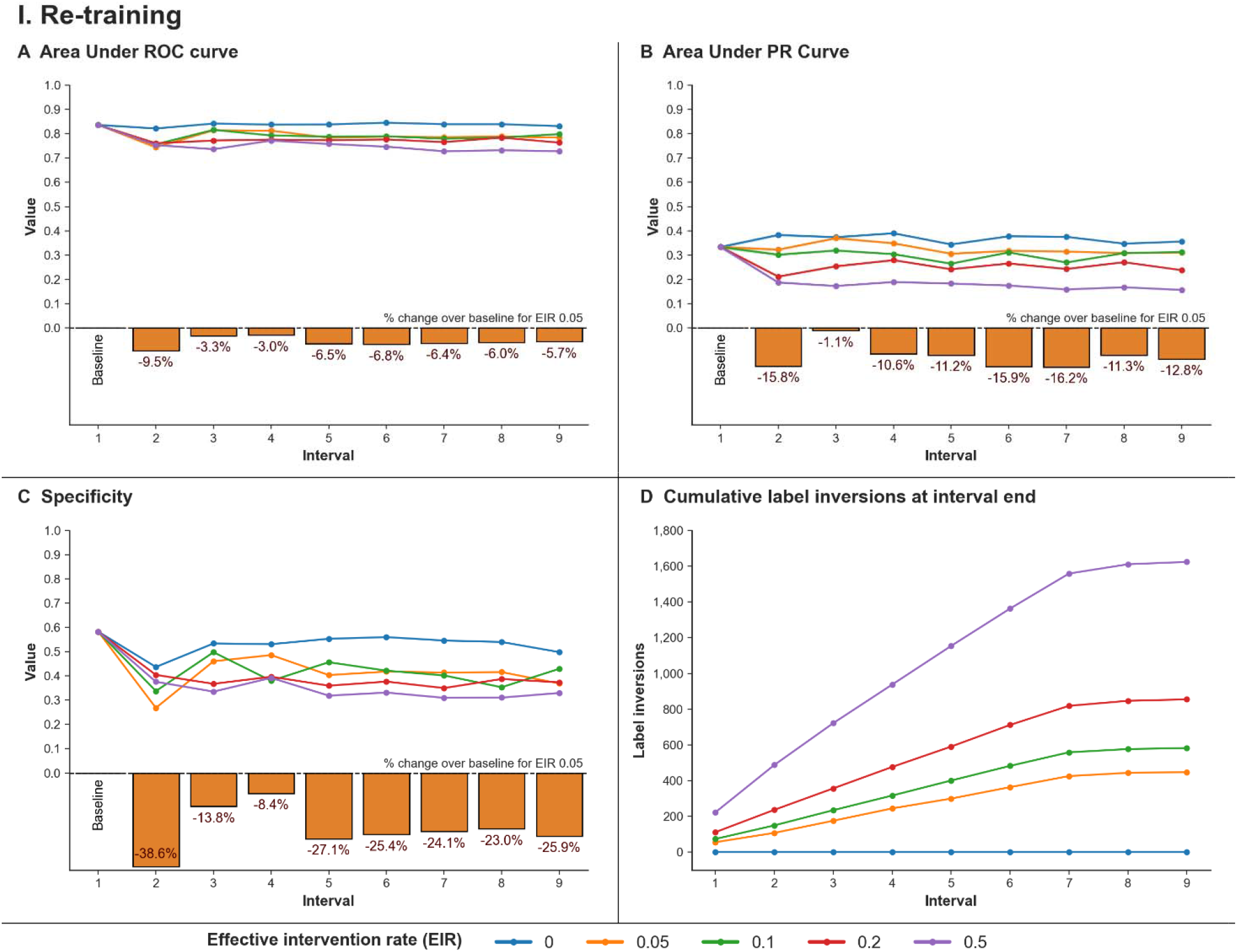
Scenario I: Re-training | Deployment and re-training of a model for prediction of post-ICU admission mortality (MIMIC IV data) *Simulation parameters – Model: XGBoost, threshold calibration: 90% sensitivity, account for effect of neglect, hyperparameter optimization performed, population class distribution maintained, no mitigation in place* Each line signifies an effective intervention rate (EIR) for a performance metric across iterations of re-training. Primary deployment of the model is at interval 1, with each subsequent interval indicating one model instance of implementation and re-training. Panels A, B, and C show changes in AUROC, AUPRC, and specificity respectively. Bar plots in panels A, B, and C show percentage difference in performance between a model developed in baseline laboratory conditions (EIR of 0), and the lowest simulated EIR of 0.05. Sensitivity is constant at 0.9 across all intervals since the model’s classification threshold is set for 90% sensitivity after each update. Panel D shows the cumulative number of label inversions leading up to an interval for an effective intervention rate. Relatively small changes are seen in AUROC, while AUPRC and specificity reduce much more dramatically depending on intervention rate and number of iterations of re-training. Drop in performance stabilizes following 2-3 iterations of re-training.

Factors that improve performance across all experiments include hyperparameter optimization, choice of XGBoost model, and either of the feature-based or patient exclusion strategies. Shuffling patient admission times to account for changes in incoming data is seen to have little effect on relevant simulations. Interestingly, model performance for prospective data is not consistent with the classification threshold derived for currently available data, with large fluctuations in both the sensitivity and specificity across each update interval. Finally, model calibration is seen to be worse after re-training with models tending to under-estimate risk **(Supplementary Figures 2, 3, 4.)** Results using Mount Sinai data show similar trends. **(Supplementary Appendix 1)**

### Scenario II – Future model development

We simulate the creation of a model for prediction of 5-day mortality post-ICU admission following the use of an initial model for prediction of 5-day AKI post-ICU admission, within the MSHS. We consider AKI linked to mortality if death occurs within 72 hours of the AKI. Utilization of the AKI prediction model as quantified by its OES leads to label inversions within the training data for the mortality prediction model.

Of a total 1,074 deaths in the training data for the mortality prediction model, 404 are linked to an AKI. Of the linked outcomes, we find 18 inversions at the lowest simulated EIR of 0.05, increasing to 185 inversions at 0.5. We observe relatively minimal changes in the AUROC over baseline regardless of EIR. A value of 0.86 possible in laboratory conditions drops by a maximum of 4.1% to 0.83 at an OES of 0.5. In comparison, AUPRC drops 10.8% (0.23 to 0.20) at an EIR of 0.05, and 28.6% (0.23 to 0.16) at an EIR of 0.5. Similarly, specificity drops 8.1% (0.63 to 0.58) at an EIR of 0.05, and 14.5% (0.63 to 0.54) at an EIR of 0.5. **(Figure 3, Supplementary Table 7)**.

**Figure 3.**
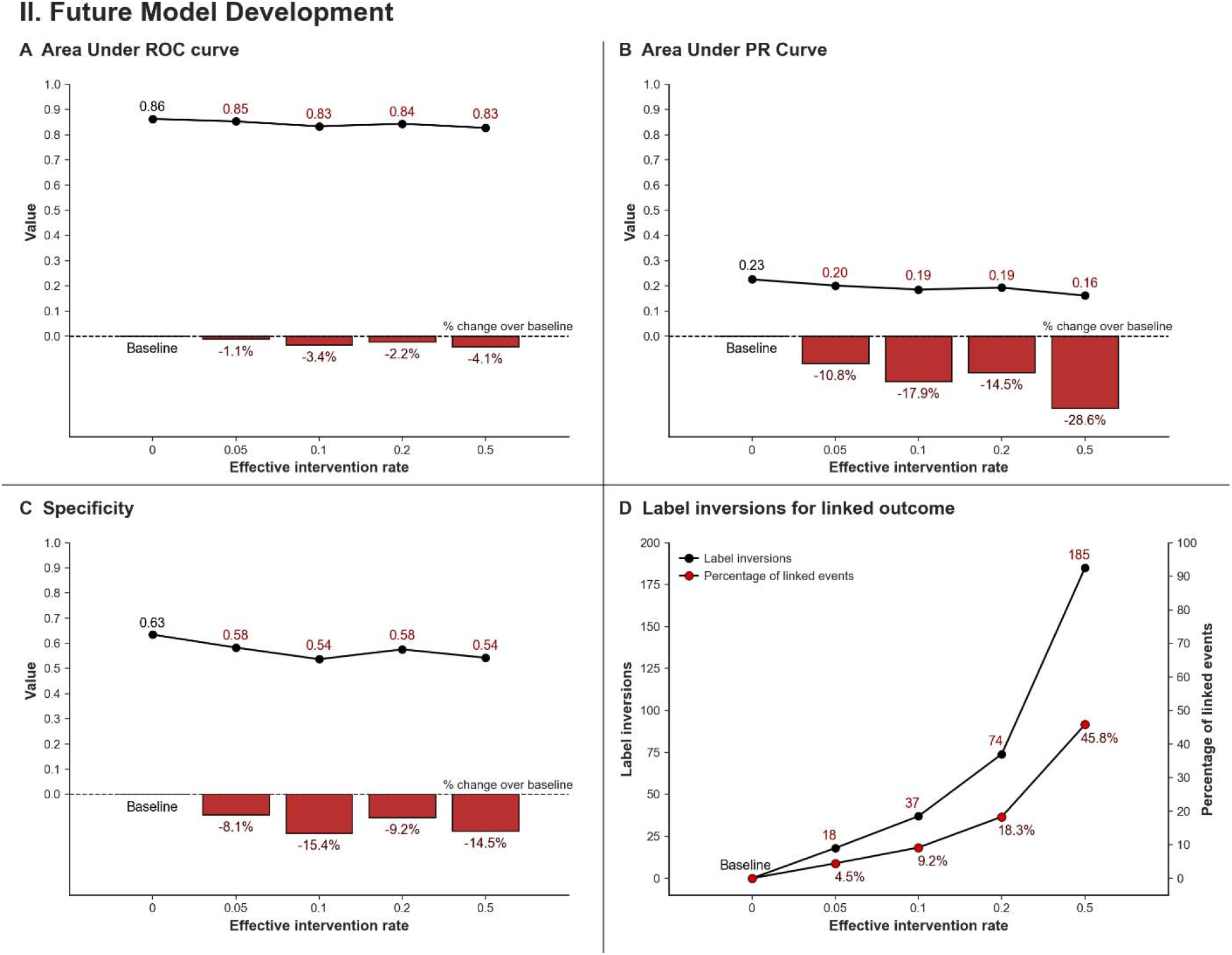
Scenario II: Future model development | Change in performance for a new post-ICU admission mortality prediction model following deployment of an initial post-ICU admission Acute Kidney Injury (AKI) prediction model (MSHS data) *Simulation parameters – Model: XGBoost, threshold calibration: 90% sensitivity, account for effect of neglect, hyperparameter optimization performed, population class distribution maintained, equal split of training and deployment data, no mitigation in place* Effective intervention rates (EIR) indicate use and effectiveness of the AKI model. AKI and mortality are considered part of the same clinical cascade if death occurs within 72 hours of the AKI. Panels A, B, and C show changes in AUROC, AUPRC, and specificity respectively with increasing effective intervention rate. Bar plots in these panels indicate percentage change in performance between a model developed in baseline laboratory conditions (EIR of 0), and each simulated EIR. Sensitivity is constant at 0.9 across all intervention rates since the new model’s classification threshold is set for 90% sensitivity. Panel D shows the number and percentage of label inversions for mortality linked to AKI due to the function of the AKI prediction model. These inversions later form part of the training dataset for the mortality prediction model. Total number of datapoints in training dataset for mortality linked to AKI: 404. Relatively small changes are seen in AUROC, with larger reductions in AUPRC and specificity given greater use and effectiveness of the AKI prediction model.

Applicable mitigations show a variable degree of correction across all performance metrics at lower intervention rates. At higher intervention rates, mitigations do not noticeably correct performance. **(Supplementary Figure 5)**

Total label inversions increase with use of a better performing model for the initial outcome, as well as leaving this model in deployment for longer. Hyperparameter optimization for the linked outcome model compensates for loss of performance to a limited extent in either case. Additionally, model calibration is seen to suffer with over-estimation of risk compared to baseline **(Supplementary Figures 4, 6, 7.)** Results using MIMIC IV data show similar trends.

### Scenario III – Multiple deployment

We simulate simultaneous deployment of a model each for prediction of 5-day mortality post-ICU admission, and 5-day AKI post-ICU admission using data from the MSHS.

Total number of invalid predictions is seen to increase exponentially with greater adherence to model predictions. Out of a total 23,053 predictions, 118 predictions for AKI, and 141 predictions for mortality are rendered invalid at an IR of 0.1. At an IR of 0.75, these values increase to 5,841 invalid predictions for AKI, and 6,962 invalid predictions for mortality.

Effective accuracy reduces slowly from baseline with increasing IR, going from 0.45 to 0.44 at an IES of 0.1 for AKI, and remaining around 0.49 for mortality. At an IR of 0.75, effective accuracy drops 25% (0.45 to 0.33) for AKI, and 28.2% (0.49 to 0.35) in the case of mortality. **(Figure 4, Supplementary Table 8)**

**Figure 4.**
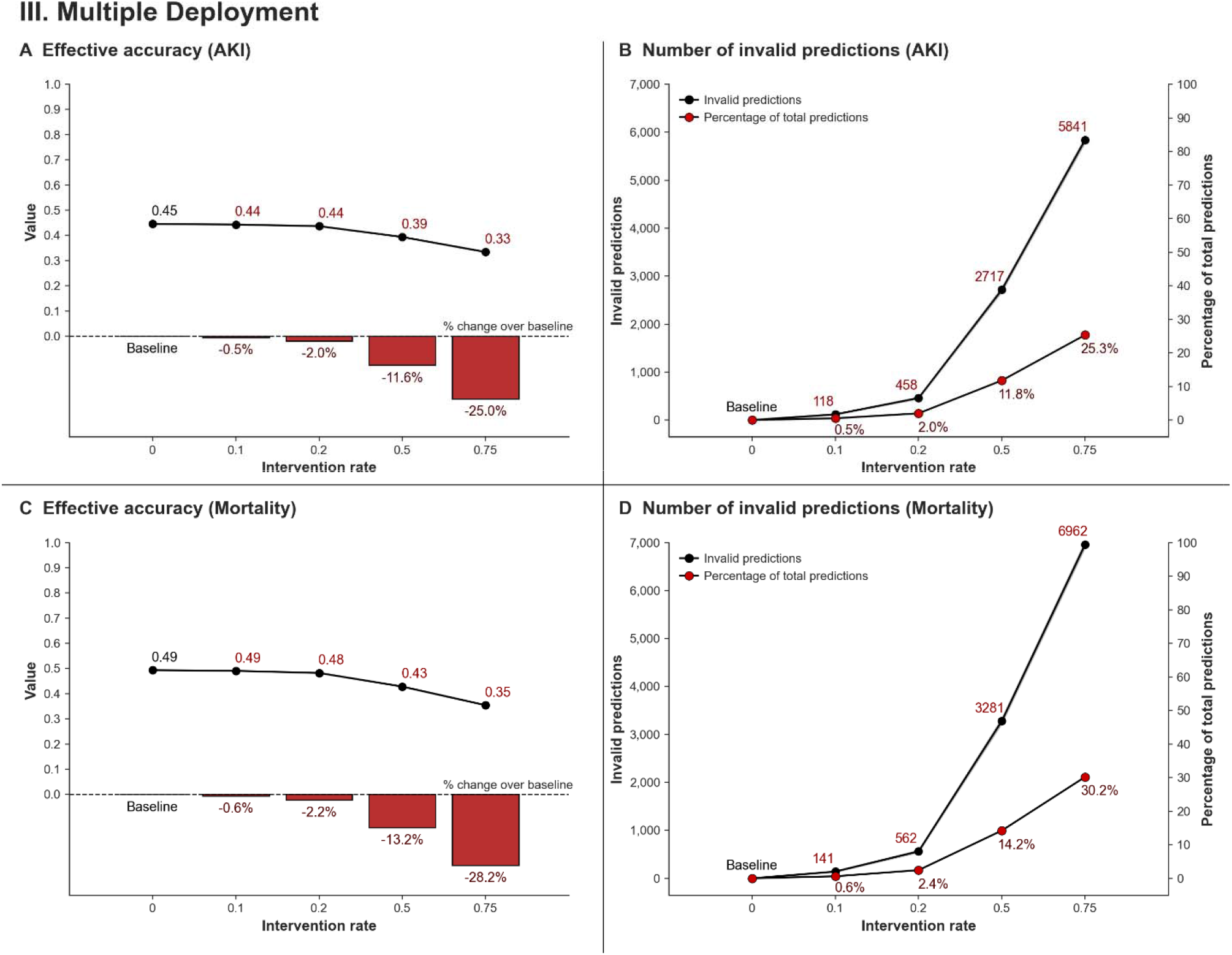
Scenario III: Multiple deployment | Simultaneous deployment of a model each for prediction of post-ICU admission Acute Kidney Injury (AKI), and post-ICU admission mortality (MSHS data) *Simulation parameters – Model: XGBoost, Model: threshold calibration: 90% sensitivity, hyperparameter optimization performed, equal split of training and deployment data, one dummy model part of system, false positive predictions are implemented* Effective Accuracy is calculated as the ratio of the number of valid predictions matching the target outcome vs the total number of predictions. Panels A and C show changes in effective accuracy for the AKI and mortality prediction models respectively with an increasing intervention rate (IR). Bar plots in panels A and C show percentage change in performance between a model developed in baseline laboratory conditions (IR of 0), and each simulated IR. Panels B and D show the number and percentage of predictions for either of the AKI and mortality outcomes made invalid by the prior implementation of a prediction from another model. Total number of predictions made by AKI and death models: 23,053. For both models, effective accuracy is seen to reduce while the number of invalid predictions is seen to increase with physician adoption of predictions.

In general, effective accuracy is seen to be affected to a much smaller extent at low intervention rates. Hyperparameter optimization is seen to have minimal impact on the total number of predictions invalidated. However, choice of the XGBoost model, *false positive* predictions affecting patient state, and additional models being active dramatically increase the overall unpredictability of the system. **(Supplementary Figures 8, 9)** Results using MIMIC IV data show similar trends **(Supplementary Appendix 1)**

## Discussion

Our findings indicate that model use precipitates interactions which damage the validity of deployed models, and of models developed in the future. We realize these findings through the creation of a simulation framework that can quantify aftereffects of model use and anticipate post-deployment issues. For the clinician, these issues may manifest as reduced specificity – entailing unnecessary investigations or procedures, reduced sensitivity – entailing neglect or underreporting of at-risk cases, or the implementation of invalid predictions leading to a combination of the above in an unpredictable manner.

Re-training has been recommended as a remedy for waning model performance due to dataset shift in recent work^14^. In Scenario I, we demonstrate that naïve re-training of a model after deployment and usage quickly and dramatically reduces its performance. The rate and extent of loss of performance is seen to be in direct proportion to the use and usefulness of the model. Previous work regarding such “*feedback loops*” is limited to consideration of unrealistic levels of physician neglect coupled to rigid adherence to erroneous model predictions^23^. As we demonstrate, re-training even in low adoption settings with no or minimal consideration of neglect is sufficient to impair a model. Equally, never updating the model may also degrade performance due to changes in incidence^24,25^, and population level or contextual changes^14,26^.

In Scenario II, we explore the effect of label inversions due to linked outcomes within clinical cascades. Simulations again show performance degradation in direct proportion to the effectiveness of the model from earlier in the cascade. While reduction in performance is lower relative to Scenario I, linkage is *additive* – the same downstream outcome (e.g., death) can form part of multiple cascades with differing upstream events (e.g., AKI, sepsis, pneumonia etc.). Further, linkage is *bi-directional* – increased monitoring for a patient at risk of a terminal cascade event may prevent an earlier linked event. Additionally, while we discuss clearly defined, overt clinical outcomes, the effect will also influence outcomes with less obvious and/or longer-term associations, with an increased magnitude when linkage occurs across longer periods of time owing to a corresponding increase in the number of label inversions. Overall, outcome linkage can result in passive, progressive dilution of label veracity within the EHR that may not be easily apparent.

ML models in healthcare may be broadly divided^27^ into *diagnostic models* which infer state at the time of data collection, and *prognostic models* which infer state at some time in the future. Interventions cannot change past diagnoses, and therefore, diagnostic models are immune to label inversions. However, deployed diagnostic models which prioritize care for sicker individuals can confound future model development by upending the relationship between severe disease presentation and eventual outcome^28,29^. Taken together, we posit that any deployed model, diagnostic or prognostic, stands to irreversibly taint present and future data for a patient for whom a prediction influences clinical care. Caution must be exercised if including such a patient within a cohort for any future prognostic model development, especially when considering their data from a period intersecting with a prior presentation-prediction cycle. While the influence of an implemented prediction may reduce with time, more work is required to identify if or when data from such patients will be valid for use again. In our experiments, enumerated mitigations **(Table 1)** do not fully correct for loss of performance. Continued dilution of EHR integrity may eventually require less efficient post-effect corrections once sufficient label inversions have taken hold. **(Supplementary Table 9)**

Label inversions leading to disruptions in the data and label relationship might appear to be similar to *Concept drift*^26,30^ – model degradation over time due to changes in a hidden context linked to the label. Literature recommends periodically re-training models to account for the change in the hidden context^26^. However, concept drift does not extend to developing models using tainted data sources; and as we demonstrate, re-training models may not be a viable solution.

In Scenario III, we consider the effect of models interacting directly at patient level. A change in the biological state of a patient either guided by, or independent of a model stands to invalidate prior predictions. The degree of change in biological state, and how this change will affect predictions from a model, cannot be easily quantified. Therefore, we believe each interaction with a healthcare provider resulting in a change in management necessitates a reissue of all applicable predictions before acting on or continuing prior model recommended paths of action. Models utilized in outpatient settings are especially important to consider in this context since issued predictions may only be updated during infrequent visits to the clinic^31^ but may influence care on a long-term basis. Within inpatient settings, getting all predictions in a batch every few hours is a potentially harmful practice which may increase the likelihood of an invalid prediction getting implemented.

Further, since simply implementing a prediction may modify biological state, considering larger intervention rates for these simulations is more reflective of reality – especially in settings with high model adoption.^32^ In comparison to models utilizing only coarse demographic or comorbidity data, commonly applicable models utilizing relatively more volatile measurements such as labs, vitals, and prior interventions are substantially more susceptible to this effect.

Cumulative effects observed across all deployment scenarios stand to increase with the number of deployed models and total deployment time – culminating not just in a loss of performance, but also confounding measurements of model performance. Owing to the permanence of data within the EHR, these effects persist despite new patients and data entering the modeling pipeline. Therefore, we *strongly* recommend immediate measures to track how and when predictions are implemented. Currently prevalent practices of opaque, unaudited, “*fire-and-forget*” deployment within the EHR, often by EHR vendors^14,33^, will result in additional burdens on healthcare systems; irreparable problems for future machine learning research and development; and impact the viability of *learning health-systems*^*34*^.

Our work is subject to certain limitations. Clinical deployment may work better with models structured as continuous risk predictors outputting calibrated probabilities instead of binary classifiers^35^. However, continuous risk models will still precipitate interactions subject to provider interpretation of low/high risk numbers, while requiring an intractably high number of simulation experiments to describe fully. Further, we only consider models working with tabular data while newer approaches use multi-modal healthcare data encompassing histopathology, imaging, and notes. However, the effects described within our experiments happen at the level of the label derived from the EHR, and models trained on multi-modal data will be subject to similar long-term implications as purely tabular models.

In conclusion, owing to the nascence of ML implementation science in healthcare coupled to a lack of long-term follow-up, current research lacks understanding of interactions between models and the EHR, and interactions between models themselves. In a model eat model world, a deployed model can confound the current operation and future development of other models, in addition to eventually rendering itself unusable.

## Data Availability

All data produced in the present study are available upon reasonable request to the authors.

## Acknowledgements

We thank Wei Guo, Lili Gai, and Eugene Fluder of the High Performance Computing group at Mount Sinai for enabling the computational infrastructure underlying these experiments. Ira Hofer receives funding from the NIH (NIH 1K01HL150318).

## Contributions

The study was designed by AV. Code for simulations was written by AV. Underlying data were collected, analyzed, and visualized by AV. First draft of the manuscript was written by AV. GNN supervised the project. AV and GNN had access to and verified the data. All authors provided feedback and approved the final draft.

## References

1. Kersting K. Machine Learning and Artificial Intelligence: Two Fellow Travelers on the Quest for Intelligent Behavior in Machines. Frontiers in Big Data 2018;1 (Specialty Grand Challenge) (In English). DOI: 10.3389/fdata.2018.00006.

2. Najafabadi MM, Villanustre F, Khoshgoftaar TM, Seliya N, Wald R, Muharemagic E. Deep learning applications and challenges in big data analytics. Journal of Big Data 2015;2(1):1. DOI: 10.1186/s40537-014-0007-7.

3. Evans RS. Electronic Health Records: Then, Now, and in the Future. Yearb Med Inform 2016;Suppl 1(Suppl 1):S48–S61. (In eng). DOI: 10.15265/IYS-2016-s006.

4. Shatte ABR, Hutchinson DM, Teague SJ. Machine learning in mental health: a scoping review of methods and applications. Psychological Medicine 2019;49(9):1426–1448. DOI: 10.1017/S0033291719000151.

5. Quer G, Arnaout R, Henne M, Arnaout R. Machine Learning and the Future of Cardiovascular Care. Journal of the American College of Cardiology 2021;77(3):300–313. DOI: 10.1016/j.jacc.2020.11.030.

6. Choy G, Khalilzadeh O, Michalski M, et al. Current Applications and Future Impact of Machine Learning in Radiology. Radiology 2018;288(2):318–328. DOI: 10.1148/radiol.2018171820.

7. Rajpurkar P, Chen E, Banerjee O, Topol EJ. AI in health and medicine. Nature Medicine 2022;28(1):31–38. DOI: 10.1038/s41591-021-01614-0.

8. Wiens J, Saria S, Sendak M, et al. Do no harm: a roadmap for responsible machine learning for health care. Nature Medicine 2019;25(9):1337–1340. DOI: 10.1038/s41591-019-0548-6.

9. Kakarmath S, Esteva A, Arnaout R, et al. Best practices for authors of healthcare-related artificial intelligence manuscripts. npj Digital Medicine 2020;3(1):134. DOI: 10.1038/s41746-020-00336-w.

10. Rajkomar A, Dean J, Kohane I. Machine Learning in Medicine. N Engl J Med 2019;380(14):1347-1358. (In eng). DOI: 10.1056/NEJMra1814259.

11. Kiani A, Uyumazturk B, Rajpurkar P, et al. Impact of a deep learning assistant on the histopathologic classification of liver cancer. NPJ Digit Med 2020;3:23. (In eng). DOI: 10.1038/s41746-020-0232-8.

12. Cohen JP, Cao T, Viviano JD, et al. Problems in the deployment of machine-learned models in health care. Canadian Medical Association Journal 2021;193(35):E1391–E1394. DOI: 10.1503/cmaj.202066.

13. Mitchell TM. The discipline of machine learning: Carnegie Mellon University, School of Computer Science, Machine Learning …, 2006.

14. Finlayson SG, Subbaswamy A, Singh K, et al. The Clinician and Dataset Shift in Artificial Intelligence. New England Journal of Medicine 2021;385(3):283–286. DOI: 10.1056/NEJMc2104626.

15. Cutillo CM, Sharma KR, Foschini L, et al. Machine intelligence in healthcare—perspectives on trustworthiness, explainability, usability, and transparency. npj Digital Medicine 2020;3(1):47. DOI: 10.1038/s41746-020-0254-2.

16. Brown JR, Rezaee ME, Marshall EJ, Matheny ME. Hospital Mortality in the United States following Acute Kidney Injury. BioMed Research International 2016;2016:4278579. DOI: 10.1155/2016/4278579.

17. Yende S, Angus DC. Long-term outcomes from sepsis. Current Infectious Disease Reports 2007;9(5):382–386. DOI: 10.1007/s11908-007-0059-3.

18. Adams R, Henry KE, Sridharan A, et al. Prospective, multi-site study of patient outcomes after implementation of the TREWS machine learning-based early warning system for sepsis. Nature Medicine 2022;28(7):1455–1460. DOI: 10.1038/s41591-022-01894-0.

19. Johnson A, Bulgarelli L, Pollard T, Horng S, Celi LA, Mark R. MIMIC-IV. PhysioNet; 2021.

20. Khwaja A. KDIGO Clinical Practice Guidelines for Acute Kidney Injury. Nephron Clinical Practice 2012;120(4):c179–c184. DOI: 10.1159/000339789.

21. Chen T, Guestrin C. XGBoost: A Scalable Tree Boosting System. Proceedings of the 22nd ACM SIGKDD International Conference on Knowledge Discovery and Data Mining. San Francisco, California, USA: Association for Computing Machinery; 2016:785–794.

22. Handelman GS, Kok HK, Chandra RV, et al. Peering into the black box of artificial intelligence: evaluation metrics of machine learning methods. American Journal of Roentgenology 2019;212(1):38–43.

23. Adam GA, Chang C-HK, Haibe-Kains B, Goldenberg A. Hidden Risks of Machine Learning Applied to Healthcare: Unintended Feedback Loops Between Models and Future Data Causing Model Degradation. In: Finale D-V, Jim F, Ken J, et al., eds. Proceedings of the 5th Machine Learning for Healthcare Conference. Proceedings of Machine Learning Research: PMLR; 2020:710--731.

24. Fleming SL, McFarlane KH, Thapa I, et al. Performance of a Commonly Used Pressure Injury Risk Model Under Changing Incidence. Jt Comm J Qual Patient Saf 2022;48(3):131–138. (In eng). DOI: 10.1016/j.jcjq.2021.10.008.

25. Lenert MC, Matheny ME, Walsh CG. Prognostic models will be victims of their own success, unless…. J Am Med Inform Assoc 2019;26(12):1645–1650. (In eng). DOI: 10.1093/jamia/ocz145.

26. Tsymbal A. The Problem of Concept Drift: Definitions and Related Work. 2004.

27. van Smeden M, Reitsma JB, Riley RD, Collins GS, Moons KGM. Clinical prediction models: diagnosis versus prognosis. Journal of Clinical Epidemiology 2021;132:142–145. DOI: 10.1016/j.jclinepi.2021.01.009.

28. Titano JJ, Badgeley M, Schefflein J, et al. Automated deep-neural-network surveillance of cranial images for acute neurologic events. Nature Medicine 2018;24(9):1337–1341. DOI: 10.1038/s41591-018-0147-y.

29. Vaid A, Johnson KW, Badgeley MA, et al. Using Deep-Learning Algorithms to Simultaneously Identify Right and Left Ventricular Dysfunction From the Electrocardiogram. JACC Cardiovasc Imaging 2022;15(3):395–410. DOI: 10.1016/j.jcmg.2021.08.004.

30. Kukar M. Drifting Concepts as Hidden Factors in Clinical Studies. Berlin, Heidelberg: Springer Berlin Heidelberg; 2003:355–364.

31. Ganguli I, Lee TH, Mehrotra A. Evidence and Implications Behind a National Decline in Primary Care Visits. Journal of General Internal Medicine 2019;34(10):2260–2263. DOI: 10.1007/s11606-019-05104-5.

32. Henry KE, Adams R, Parent C, et al. Factors driving provider adoption of the TREWS machine learning-based early warning system and its effects on sepsis treatment timing. Nature Medicine 2022;28(7):1447–1454. DOI: 10.1038/s41591-022-01895-z.

33. Wong A, Otles E, Donnelly JP, et al. External Validation of a Widely Implemented Proprietary Sepsis Prediction Model in Hospitalized Patients. JAMA Internal Medicine 2021;181(8):1065–1070. DOI: 10.1001/jamainternmed.2021.2626.

34. Menear M, Blanchette M-A, Demers-Payette O, Roy D. A framework for value-creating learning health systems. Health Research Policy and Systems 2019;17(1):79. DOI: 10.1186/s12961-019-0477-3.

35. Van Calster B, Vickers AJ. Calibration of risk prediction models: impact on decision-analytic performance. Medical decision making 2015;35(2):162–169.

36. He H, Garcia EA. Learning from Imbalanced Data. IEEE Transactions on Knowledge and Data Engineering 2009;21(9):1263–1284. DOI: 10.1109/TKDE.2008.239.

37. Price WN. Big data and black-box medical algorithms. Sci Transl Med 2018;10(471) (In eng). DOI: 10.1126/scitranslmed.aao5333.

